# A systematic literature review of individual-level psychological and behavioral responses to the health information of COVID-19 from social media and legacy media

**DOI:** 10.1101/2021.12.14.21267757

**Authors:** Wei Zhai, Jian Bian, Jing Zhang, Xueyin Bai

**Author notes:** Corresponding author: Dr. Wei Zhai.

## Abstract

Covid-19 has been recognized as a terrifying global health threat since its detection, with far-reaching consequences that are unprecedented in the modern era. Since the outbreak of the pandemic, social media and legacy media have collectively delivered health information related to COVID-19 to the public as a catalyst to community perception of risk. However, the existing literature exhibits different viewpoints toward the role of social media and legacy media in disseminating health information of COVID-19. In this regard, this article conducted a systematic literature review to provide an overview of the current state of research concerning individuals-level psychological and behavioral response to COVID-19 related information from different sources, as well as presents the challenges and future research directions.

## 1. Introduction

The 2020 novel coronavirus pandemic has caused a massive impact on the world, which is not only a public health crisis but also an unprecedented challenge for economic development and people’s daily lives (Muhammad et al., 2020; Zhai and Peng, 2021). According to the World Health Organization (WHO), as of November 23, 2021, over 258 million positive cases of COVID-19 globally have been confirmed, including 5.16 million deaths. In particular, the Delta variant, with its highly infectious nature, is now exacerbating the global transmission of new coronaviruses. To combat the pandemic, many countries have implemented different intervention strategies, such as restriction of mass gatherings, reduced frequency of public transportation, lockdown of city, etc. (Musselwhite, Avineri and Susilo, 2020; Brauner et al., 2021; Zhai and Yue, 2021). The effectiveness of government interventions heavily depends on the general public’s support for, compliance to, and trust in the policies (Liu et al., 2021; Zhai et al., 2021a; Zhai et al., 2021b; Zhai et al., 2021c). In this process, it is the social media and legacy media working as an intermediary bridge between authorities (government/professional medical institutions) and the public, regulating and influencing public awareness and behaviors (Krawczyk et al., 2021; Liu et al., 2020; Zhou et al., 2020). For instance, in most cases, people would internalize the received information and then adopt protective actions, such as home quarantine, social distancing, washing hands with alcohol, wearing masks, and vaccination, to slow virus transmission (Wang et al., 2021; Freeman and Eykelbosh, 2020; Fu and Zhai, 2021). In addition, many individuals seek to receive timely information (Van et al.,2021), or keep social connections with important ones (Minh et al., 2021) to ease anxiety from the infection risk.

Even though social media have played an important role in disseminating health-protection guidelines and delivering the latest information on pandemics to the public, the impacts of social media communications on the pandemic are mixed. On the one hand, social media has the advantages of transparency such as the huge number of accessible channels, various types of resources, up-to-date information, and integrating hyperlinks (Patrick et al., 2019; Jin et al., 2019; McInnes and Hornmoen, et al., 2018). To this end, social media users can easily get access to, share, and generate health-related content on social media (Chen and Wang, 2021). In the meantime, social media users also can enhance community cohesion and help people gain emotional resonance and consensus for action during the pandemic. On the other hand, in today’s hyper-networked communication environment, the information is flowing through every social media user via the huge and complicated network with uncertainties, instead of being filtered by gatekeepers of information verification, for example, on legacy media (Shoemaker and Vos, 2009). Thus, social media users are quite prone to problems such as infodemic (WHO, 2020), referring to widespread misinformation and information overload (Pulido et al., 2020; Bode et al., 2021).

The individuals’ responses to the information from legacy media (i.e., radio, television, newspaper, etc.) are also two-sided. The gatekeeping role of mainstream media is quite important because it can filter out useless information, especially from the noisy social media environment, help the general public conduct fact-checking, and alleviate the psychological fear and anxiety of individuals (Ophir & Jamieson, 2020). In addition, traditional media can be effective in providing credible information in an effort to increase public awareness of prevention and intervention strategies (Weick,1988; Ophir., 2019; Bao et al., 2020). However, the existing literature indicates that the news coverage of COVID-19 shows signs of being politicized and used by politicians to serve particular ideological interests (Earnshaw et al., 2020). For instance, in a highly partisan environment such as the United States, political bias has damaged the delivery of health messaging via legacy media due to the contested trust of the science in left-leaning and right-leaning media (Zhao et al., 2020).

Therefore, social media and legacy media have made significant impacts on people’s risk perceptions, emotions, and preventive behaviors during the pandemic (Tsai et al., 2020). The main objective of this article is to review the existing literature so as to understand how individuals respond to the health information pertaining to COVID-19 from both social media and legacy media. For this purpose, a methodological approach based on a systematic literature review is applied, which provides an overview of the current state of research, as well as presents the challenges and future research directions. Specifically, this research aims to answer the following research questions.

- Q1. How many studies have studied the individuals’ responses to health information of COVID-19?
- Q2. Which disciplines and journals lead this research topic?
- Q3. What are the differences between social media and legacy media in terms of affecting individuals’ responses during the pandemic?
- Q4. What are the main concerns addressed by the researchers regarding individuals’ psychological and behavioral responses?
- Q5. What are challenges that have been identified by researchers in leveraging social media and legacy media to contain the viral spread?
- Q6. What are future agendas for researchers and practitioners?

To address Q1 and Q2, the articles published in the journal are identified in Section 2. Regarding Q3, Q4, and Q5, the scopes of the existing studies are analyzed, as well as the cases and the applied techniques applied by researchers in Section 3 and 4. Finally, to answer Q6, we will propose several promising agendas for future studies in Section 5. Finally, Section 6 will conclude the research.

## 2. Research design and methods

### 2.1 Searching protocol

This study applied two strategies to review the current state of research about the relationship between media exposure, health-protective behaviors, and viral spread amid COVID-19. The first strategy is to identify all English-language studies that were published since 2020 from Web of Science and Google Scholar databases. A combination of the following keywords was used to search for titles and abstracts: “media” OR “response”. Figure 1 shows two types of search categories with different keywords in each category of search terms. The category “media” can be specified as media-related behavior (“media coverage” / “media use”) and media type (“social media” / “legacy media”). The category “response” mainly includes psychological responses, for instance, including “risk perception”, “health purposes” and “mental health consequences” and behavioral responses such as “behavioral response”, “travel” / “stay at home” / “hand wash”, etc. Each keyword in Figure 1 relates to the pandemic using “AND” and the keyword “COVID-19” in every search. The second strategy was collecting articles from the reference list of existing literature.

**Figure 1.**
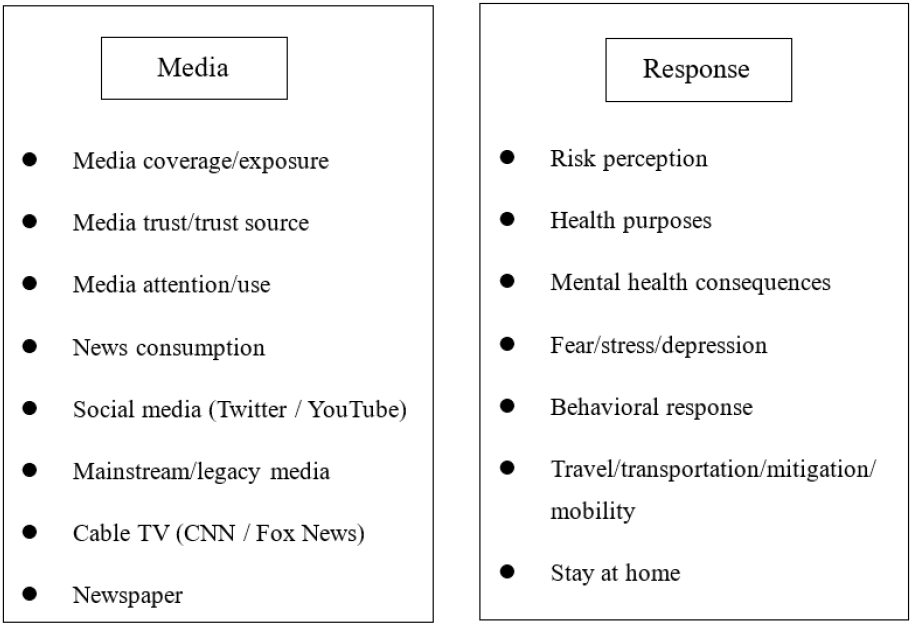
Keywords under the category “media” and “response**”**

### 2.2 Screening protocol

As indicated in Figure 2, the screening process was developed from the PRISMA (Preferred Reporting Items for Systematic Reviews and Meta-Analyses) flow diagram with four steps: identification, screening, eligibility, and inclusion (Moher et al., 2009; Lu et al., 2019). The step of identification selected all articles that explored the impact of the media or investigated the public’s health and behavioral responses during the pandemic. The step of screening filtered out the studies that ignored the connection between media coverage and health responses during the pandemic. Articles published before 2020 were excluded. The step of eligibility excluded literature review articles to ensure that selected research articles are evidence-based studies. The step of inclusion further excluded non-peer-review journal articles and articles with duplicated research themes and findings. As of September 2021, there were 33 articles from Web of Science, 8 articles from Google Scholar, and 14 articles identified from reference lists of those articles, resulting in a total of 55 articles (Appendix 1).

**Figure 2.**
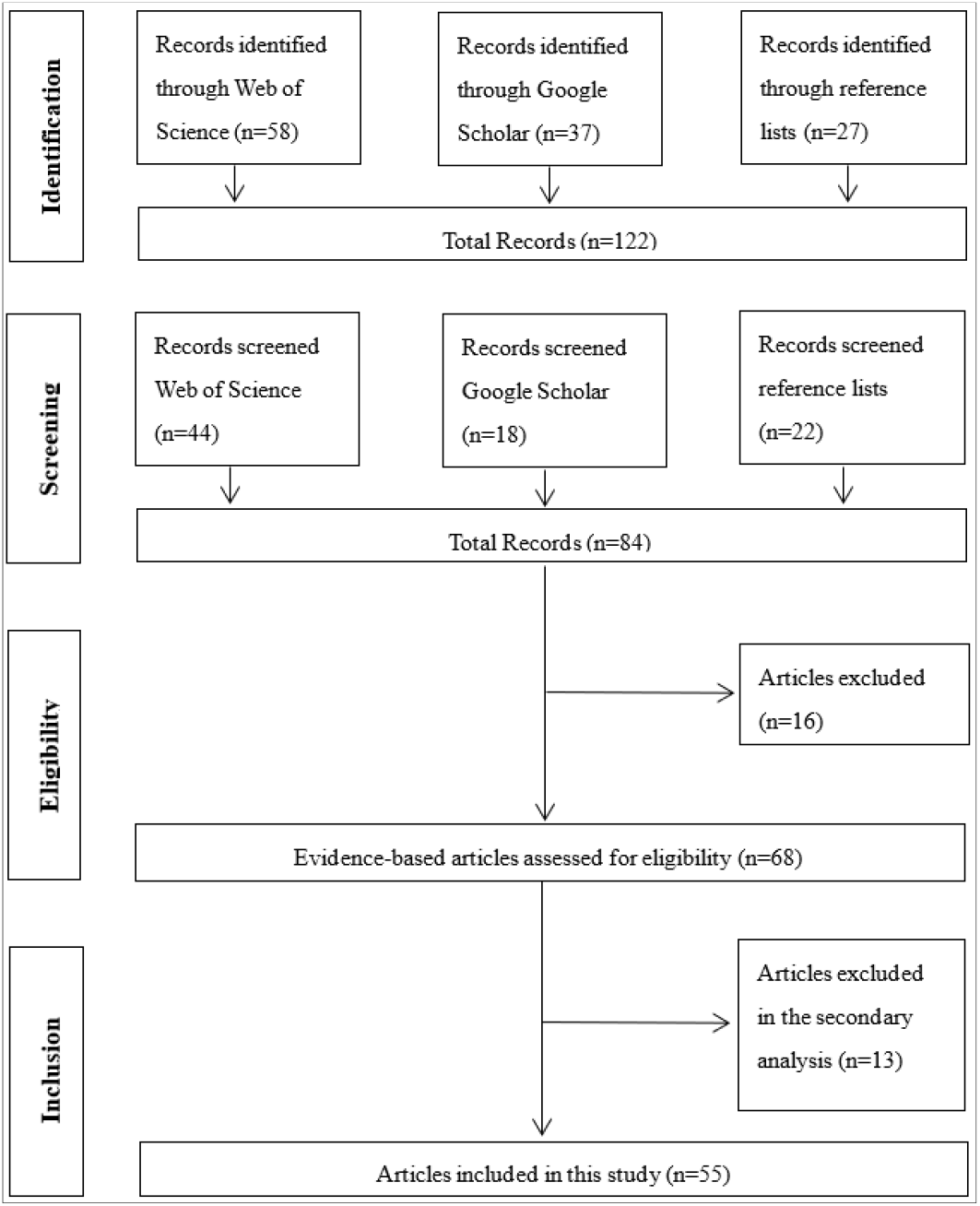
Flowchart of systematic literature review process and number of included and excluded papers in each step

## 3. Review results

### 3.1 Type of documents

#### Journal impact

Journal Citation Report (JCR) Impact Factor (IF) reflects the average number of citations, which is a measure of the quality of a journal to some extent. Table 1 shows the impact factor and the quartile of the recorded articles. Clearly, the majority of selected papers were published in journals ranked in Q1 and Q2. Especially noteworthy is that *Journal of Medical Internet Research*, as a Q1 journal, has published 7 articles on this topic, followed by *Health Communication, Social Media + Society* and *Mathematical Biosciences and Engineering*.

**Table 1.** A summary of journals

| Journals | Count | Quartile | IF |
| --- | --- | --- | --- |
| Journal of Medical Internet Research | 7 | Q1 | 5.428 |
| Health Communication | 3 | Q2 | 3.198 |
| Mathematical Biosciences and Engineering | 3 | Q3 | 2.08 |
| Social Media + Society | 3 | Q1 | 4.249 |
| BMJ Global Health | 2 | Q1 | 5.558 |
| Technological Forecasting and Social Change | 2 | Q1 | 8.593 |
| Plos One | 2 | Q2 | 3.24 |
| Journal of Health Communication | 2 | Q2 | 2.781 |
| JMIR Public Health and Surveillance | 2 | Q1 | 4.112 |
| EPJ Data Science | 2 | Q1 | 3.184 |
| Others | 29 |  |  |

#### Discipline

Table 2 indicates the disciplines that are related to at least two articles. It is noticeable that the articles are published in multiple disciplines. *Communication*, as a subject correlated with delivering the message to different people, holds the lead position on this research topic. Disciplines like *Public Health, Computer Science, Information Science, Management, Mathematics, Behavioral Sciences*, and so on, all put their lens to concentrate on a different aspect of this topic. With more multidiscipline cooperation, there will be more practicable suggestions to mitigate the global crisis made by this novel coronavirus.

**Table 2.** A summary of disciplines

| Discipline | Numbers |
| --- | --- |
| Communication | 18 |
| Public Health | 6 |
| Computer Science | 4 |
| Information Science | 4 |
| Mathematics | 4 |
| Management | 3 |
| Psychology | 3 |
| Behavioral Science | 2 |
| Journalism | 2 |
| Media | 2 |
| Medical | 2 |
| Medicine | 2 |
| Political Science | 2 |
| Tourism | 2 |

**Table 3.** A summary of countries (August 24, 2021)

| Countries | Numbers | Confirmed Cases | Deaths |
| --- | --- | --- | --- |
| United States | 18 | 37,939,641 | 629,411 |
| China | 11 | 106,812 | 4,848 |
| Global | 11 | 212,717,174 | 4,445,450 |
| United Kingdom | 9 | 6,555,419 | 132,000 |
| Italy | 6 | 4,488,779 | 128,795 |
| Germany | 4 | 3,881,633 | 92,028 |
| France | 4 | 6,708,163 | 113,857 |
| Spain | 4 | 4,794,352 | 83,337 |
| Canada | 2 | 1,481,438 | 26,780 |
| Saudi Arabic | 2 | 541,994 | 8,481 |
| South Korea | 2 | 239,287 | 2,228 |
| Nigeria | 2 | 187,588 | 2,276 |

Note: Some studies conducted a case study in different countries so that the total number is greater than the number of articles.

#### Country

The total number of people infected in the United States was 37,939,641 till August 24, 2021, ranking the first in the world. Therefore, it is not surprising that the case studies of the United States take the lead position on this theme. China keeps a low infectious rate compared to other countries, but scholars are very active on this topic. It is worth noting that 11 articles investigated the global impacts of social media or legacy media on viral transmission, indicating that understanding this issue is also a global challenge, rather than an area-focused topic.

### 3.2 Social media and legacy media

While individuals’ responses to health information are clearly influenced by the media platform, the literature found that social media and legacy media exhibit different patterns.

Social media is known for its nature of real-time. Different from legacy media that relies on editors or agencies to publish news, every social media user can publish information online without restrictions. There are seven studies that show a strong correlation between social media activities about COVID-19 and viral transmission. For instance, Bisanzio et al. (2020) and Cui and Kertész (2021) demonstrated that social media (e.g., Twitter, Weibo) can be used to predict the spatial and temporal transmission of infectious diseases in pandemic times. Younis et al.(2020) confirmed that the utility of social media can be used as an epidemiological tool in the assessment of protection measures at the beginning of the pandemic. Amara (2021) traced COVID-19-related topics in 7 languages on Facebook and then found that the topics are consistent with the pandemic evolution. Furthermore, Haouari et al. (2020) found that the early reported cases of COVID-19 were correlated with the heated discussion of topics on Twitter when the virus appeared in the Arab world. Zhang et al. (2021) found significant associations between human mobility and social media topics concerning COVID-19 dissemination, and societal interventions from February 15 to June 14, 2020, in Denmark and Sweden. Based on statistics from the multilingual COVID-19 dataset, Chen et al. (2020) indicated that Twitter activity responds and reacts to COVID-19-related events across the world.

Moreover, four studies illustrated that proper use of social media can be seen as an effective tool to curb the spread of COVID-19. Sleigh (2021) found that social media influencers from Twitter can play a crucial role in controlling the dissemination of health risk messages and promoting solidarity. Besides, Niburski and Niburski (2020) found that after Donald Trump’s Twitter posts about COVID-19, Google searches and Amazon purchases about the protective supplies were very likely to increase. Wang et al. (2021b) claimed that public health agencies and the government could take full use of the COVID-19 related information from Twitter to enact effective measures to curb the spread of the virus. Raamkumar et al. (2020) developed deep learning-based text classifiers to characterize health behaviors from social media and suggested that public health authorities could use the data to enact more effective measures.

The information from legacy media plays a vital role, especially during the early outbreak of the pandemic, offering first-hand virus-related information (Olagoke et al., 2020; Abdekhoda et al., 2021). Also, public opinion is mainly influenced by media coverage. There are seven studies examining the effect of legacy media on the public response to COVID-19. For example, Melki (2020) found that people often rely more on legacy media, especially on television, than social media during the early stage of coronavirus disease. Soroya et al. (2021) found that legacy media were more preferred than social media, and people were prone to getting news from traditional sources such as newspapers, radios, cable television, as well as online resources such as official websites. Likewise, Niu et al. (2021) found that there is a strong relationship between legacy media exposure for COVID-19-related news and preventive behaviors. They also found that high-risk groups are more likely to trust the information from TV for COVID-19-related news. The outlets of legacy media are often accompanied by media frames, Ophir et al. (2021) found that different media frames, which focused on social, political, and economic issues, were associated with increased mobility. Although people like to seek news from traditional media, Liu et al. (2020) found that legacy media news reports in China lagged behind the spread of COVID-19 in the early stage of the outbreak. Zhou et al. (2020), for example, used cross-correlation analysis to find the association between media coverage and infection cases. Chang et al. (2020) found that as the dissemination rate of health information from legacy media decreases, the confirmed cases would increase significantly.

There are four studies that examined the combined influences of social media, legacy media, and other communication tools. Krawczyk (2021) found that the quality and quantity of information from various communication channels, including social media and legacy media platforms, collectively influenced individuals’ understanding of public health measures. Melki (2020) found that those who would like to enhance protective behaviors were highly exposed to both legacy media and social media. Gozzi et al. (2020) found that user activity on Reddit and active search on Wikipedia pages about COVID-19 are primarily driven by media coverage. Lee and You (2021) found that in addition to social media or legacy media, the warning text messages were also proved to be an effective way for enhancing protective behaviors in the pandemic.

### 3.3 Psychological responses

Social media and legacy media will directly impact individuals’ psychological responses to the pandemic. In the early stage of COVID-19, exploding untrustworthy information can cause social media fatigue, thereby reducing personal information processing capabilities and increasing cognitive overload. First, two studies investigated how media outlets constructed people’s perceptions of COVID-19. Bekalu et al. (2021) found that exposure to different media platforms is correlated with different perceptions about the mortality of the infection, while the perception of virus threat is related to the high level of trust in the healthcare system and government. By analyzing the effectiveness of YouTube animated cartoons on health behaviors, Onuora (2021) determined that the perception of the seriousness of COVID-19 was the most prominent factor that influence individuals’ behaviors.

Second, the fears and uncertainty accompanied by the virus, along with anxiety and stress, have exacerbated mental health issues to some degrees throughout society (Su et al., 2021). Due to the lack of effective treatments, Ngien and Jiang (2021) found that social media affects stress through fatalism, which means human health is pre-determined by fate, chance, luck, or God, and beyond ability. Krawczyk et al. (2021) analyzed the topics from COVID-19 news and found that articles that mentioned fear, crisis, and death accounted for 16% of COVID-19 online news. Bendau et al. (2020) conducted a web-based survey in Germany and found that media usage was positively correlated with COVID-19 fear and anxiety. After a cross-sectional survey of adults living in Lebanon, Melki (2020) found that people who show a higher fear level were more likely to be exposed to a large volume of information from TV. Also, those who obtained information frequently from both TV and social media are more likely to exhibit a high level of anxiety.

Third, there is a link between conspiracy beliefs and the frequent use of social media. Chadwick et al. (2021) found that there is a strong correlation between medical conspiracy beliefs and reluctance to engage in health protection behaviors. Earnshaw et al. (2020) found that 33% of participants in their survey believed one or more conspiracies of COVID-19. Allington et al. (2020) found that the more conspiracy social media users believe, the less protective behaviors will be done. Tang et al. (2021) found that government social media promote users’ information security behavior towards COVID-19 scams.

### 3.4 Behavioral responses

Evidence from CDC shows that COVID-19 can be largely mitigated by protective behaviors. Due to the lack of efficient medication and vaccination in the early outbreak of the pandemic, Perrotta (2021) argued that protective behaviors, which are influenced by media platforms or agencies, are key to curbing the spread of the virus. Feng et al. (2020) developed a COVID-19 epidemic model based on the efficacy of media coverage to provide a possible intervention to reduce COVID-19 infection. Gozzi et al. (2020) discovered that media coverage has the potential to influence viral transmission by triggering behavioral changes. Specifically, three types of behavioral responses can be documented.

First, wearing face masks might be seen as a case of bottom-up behavioral change in Western civilizations. Perrotta et al. (2021) found that individuals’ risk perceptions of COVID-19 from media coverage were comparatively low, while they saw a rapid increase in mask use when mask-wearing was not yet mandated. The resistance of behavioral change still exists in the context of preventing and slowing the spread of the virus, evidenced by stories of individuals’ refusal to wear masks or follow social distancing guidelines that have circulated in the news during the pandemic (Ball and Wozniak, 2021). But in different countries, wearing face masks varies greatly, ranging from about 7% in the Netherlands to about 60% in Italy (Perrotta et al., 2021).

Second, information from media coverage has a major impact on raising the risk awareness of potential travelers during the pandemic, thereby affecting people’s mobility. Chemli et al. (2020) discovered that the trustworthy media coverage had a positive effect on potential travelers’ awareness of the risk of COVID-19 infection (e.g., outbound tourists’ awareness) at a national level. By collecting the data from different countries, Perrota et al. (2021) found that reduced transportation usage was found to be the most frequently reported behavior based on a survey with Facebook users, with estimates ranging from 67% in the Netherlands to 82% in Spain. Ophir et al. (2021) found that different media frames were associated with mobility changes in Italy. Huang et al. (2020) found that Twitter activities may be used to signal mobility dynamics and evaluate the effectiveness of containment measures during the pandemic. Liu et al. (2021) found an inverse U-shaped curvature in the effect of media coverage on the spread of COVID-19 in China and then mediated by intra-and inter-provincial population mobility. Melki et al. (2020) demonstrated that media exposure to COVID-19 news positively relates to people’s compliance with prevention measures. Wu and Shen (2021) found that using central government media and WeChat is associated with higher levels of compliance with health behaviors in China, but using local media and Weibo is linked to lower levels of compliance.

Third, getting a vaccine or not may also be influenced by the individuals’ exposure to information from different media sources. For instance, Chan et al. (2020) found that the more discussion of vaccination on social media, the higher vaccine willingness, and increased vaccination rate. Chadwick et al. (2021) argued that people who gain information from more different types of media are more likely to be encouraged to get a vaccine. Earnshaw et al. (2020) found that individuals, who believed conspiracies about the information from legacy media, are less likely to get a vaccine.

## 4. Challenges

### 4.1 Partisan effects

During the pandemic, some health information from legacy media was not effective in containing misleading information due to the highly partisan environment. By drawing attention to the partisan effect during the early outbreak of COVID-19, Motta et al. (2020) examined two types of media sources, one for mainstream news like New York Times or USA Today, and another for conservative media outlets such as Fox News. The results indicate that, compared to left-leaning media, right-leaning media discussed more misinformation about COVID-19. Moreover, people who obtained information from right-leaning media in the early stage were more inclined to believe the untruths than those who received information from left-leaning media. The COVID-19-related news from legacy media like the New York Times and Global Times, according to Abbas (2020), politicized the epidemic to serve the interests and ideologies of the governments from where they come from. Right-leaning and left-leaning politicians, as well as the media, have spread polarized information about COVID-19 through media. Krawczyk et al. (2021) set a sentiment analysis for media coverage of COVID-19 and found that the pandemic contents cannot be simply categorized as negatively nor positively polarized, 16% of COVID-19 related news could be classified as highly polarized.

Even though legacy media plays a crucial role in disseminating health information, several studies have shown that misinformation from different partisan media imposed a huge impact on individuals’ protective behaviors. Allcott et al. (2020) and Ananyev et al. (2021) found that there are significant conflicts between Republicans and Democrats in prevention behaviors like social distancing and wearing masks. Simonov et al. (2020) discovered that for every 10% increase in Fox News cable TV ratings, the tendency to stay at home decreased by 1.3 percentage points. Slanted information, according to Zhao et al. (2020), can impair containment efforts by influencing people’s behavior. In the United States, right-leaning media followers engaged in fewer COVID-19-related preventive actions and were involved in more risky behavior than left-leaning media followers. For example, Bekalu et al. (2021) discovered that republicans who are exposed to conservative media outlets had low trust in the reliability of scientists’ information, while democrats have high confidence in science. Hence, it is especially essential for authorities to spread unbiased information without partisan splits during a health crisis. However, Liu et al. (2020) found that people are more willing to trust the information from the government in China, irrespective of social media platforms or state-controlled media, which is directly related to their compliance with health behaviors.

### 4.2 Infodemics

Not only is the coronavirus causing chaos, but COVID-19-related infodemics, which are transmitted by individuals or groups with various political or economic motivations. Infodemics, according to Su et al. (2021), are the deliberate spread of misinformation and disinformation by the mass media, particularly on social media platforms. Wang et al. (2021a) found that that the number of users tweeting about COVID-19 health beliefs was amplifying in an epidemic manner and could partially intensify the infodemic. According to Motta et al. (2020), social media users are frequently exposed to misinformation and are prone to believe that public health officials overestimated the seriousness of the pandemic. People were often influenced by a variety of factors to spread misleading information COVID-19. Apuke and Omar (2021), for example, discovered that altruism motivation was the most important factor that determines COVID-19 fake news sharing. Some scholars have also proven that political ideologies, racial discrimination, and stigmatization in the media promoted the dissemination of misinformation in the early phases of the global epidemic (Wen et al., 2020; Motta et al., 2020; Cho et al., 2021).

Some other studies focus on modeling the predictors of fake news transmission among social media users such as altruism, unawareness, peer pressure, and also investigate how ideologies interfere with the spread of information (Apuke and Omar, 2021). Bode and Vraga (2021) found that correction behaviors on social media are common and even across the partisan divides and those with better education are more likely to engage in correction misinformation. After investigating the usefulness and reliability of the most popular YouTube videos, Li et al. (2020) found that approximately 33% of the most popular COVID-19-related YouTube videos included incorrect facts. People who are driven by self-promotion and entertainment, as well as those with poor self-regulation, are more likely to disseminate unconfirmed information, according to Islam et al. (2020). To mitigate the misinformation during the COVID-19 pandemic, Shirish (2021) believed that economic and media freedom is the most important.

Information overload can also be seen as one representative type of infodemics. Social media exposure has a close relationship with information overload (Soroya et al., 2021; Olagoke et al., 2020). According to Gao et al. (2020), there is a substantial prevalence of mental health issues, which is strongly correlated with frequent social media exposure during the COVID-19. In addition, three studies found that information and communication overload is the most important factors that lead to social media fatigue about COVID-19 information. For example, Bendau et al. (2021) found that the usage of social media was associated with psychological strain such as a higher degree of unspecific anxiety and depression about COVID-19. Ball and Wozniak (2021) found differences in information fatigue based on political affiliation such that republicans and independents scored significantly higher than democrats in information fatigue. Solomon et al. (2021) discovered that increased television news watching was related to a heightened risk for PTSD of COVID-19 infections, especially for vulnerable people.

The lack of mental health services, the overload of media outlets, and the uncertainty about the virus are all making the consequences of infodemics even worse. Su (2021) argued that media organizations paid little attention to how coverage would affect people’s mental health. Therefore, there is an urgent need for media agencies to develop a fact-based, person-centered, and collaborative response to COVID-19. Mauri-Ríos (2021) suggested that it is essential for journalists to receive clear guidelines describing how to deal with both current and future coverage of COVID-19, or other health crises. Gozzi et al. (2020) found that public health authorities need to explore more possibilities of communication channels, such as setting up accounts on social media platforms.

### 4.3 Social equality

The stigmatization of social equity during the pandemic has been evidenced. First, the mental health of ethnic/racial minorities might also be affected as a result of improper and biased media coverage. Specifically, Wen et al. (2020) suggested that racial discrimination stemming from the pandemic should be treated as a public health crisis. Tsai et al. (2020) found that people who rely more on legacy media and higher levels of trust on social media were positively related to prejudice against Asians. On the contrary, consuming news from left-leaning media and non-partisan media was linked to prejudicial attitudes toward Asians. Cho et al. (2021) found that racial prejudice is the main factor, while the use of social media and partisan cable television further exacerbated the prejudice. Although African Americans had lower vaccination intention than other groups, according to Woko et al. (2020), belief in information sources alone cannot explain the association between race and vaccination willingness. Second, gender and age also influenced the media exposure to various COVID-19 related information. Melki et al. (2020) discovered that women are more adept at mediating perceived knowledge and fear. Perrotta et al. (2021) determined that women showed higher threat perceptions than men and were more likely to have lower confidence in the health care system on social media than the man during the pandemic. Also, COVID-19 infection placed elderly people at the highest risk of serious consequences (Perrotta et al., 2021). Therefore, public health organizations should effectively use strategic health messaging tools to enhance awareness among people from all socioeconomic and demographic backgrounds.

## 5. Future directions

### 5.1 Reducing information overload

The existing studies indicated that multiple information sources are significantly associated with information overload, which can further result in various psychological and behavioral responses, such as information seeking (Soroya et al., 2021), and information anxiety (Ball and Wozniak, 2021). Thus, to reduce the effects of information overload, future studies should consider the relationship between information anxiety, familiarity with social media channels, and the technostress of social media use (Ahmad and Amin, 2012). Furthermore, the way social media messages are organized (e.g., images, language style) is also worth investigating in future studies. In addition, how personal attributes and motivational factors influence social media fatigue also remains to be examined in further research, such as a focus on the influential relationship between different purposes of social media use (entertainment, self-promotion, information seeking, information sharing) and media fatigue (Islam et al., 2020). It is also essential to explore how the public responds to the dissemination of misinformation after ideological involvement by exploring how to correct and offer more recommendations on self-regulation (Bode et al., 2021). From the perspective of the research method, in addition to using interviews and retrospective surveys, future research can conduct control experiments to test whether the factors of messages (e.g., narrative perspective, references, emotional stance) influence reactance toward COVID-19 messaging (Soroya et al., 2021).

### 5.2 Applying Artificial Intelligence

Artificial intelligence (AI) can analyze public attitudes in real-time and track changing public sentiments, using Natural Language Processing (NLP) and Machine Learning (ML) (Khan et al., 2020). Sentiment analysis uses computational methods to identify points of view in text, audio, and/or video to determine the author’s attitudes toward the topic under discussion during the pandemic (Hussain and Sheikh, 2021), offering the opportunity to detect the text polarity and identify trends in public opinion. For instance, in future studies, sentiment analysis can be performed to assess public confidence in ongoing vaccine trials in real-time and tap into constructive opinions. Thus, the AI approach can promote participatory dialogue on vaccine-related issues and other risk management topics (Ahuja et al., 2020; Hussain et al., 2021). Second, AI technology can also help disseminate credible information globally and reduce the spread of disinformation about COVID-19. Future studies should focus on the role of AI technology in fact-checking media coverage and detect the sources of the viral spread of fake news, including deceptive and sarcastic language sometimes used by vaccine skeptics. Also, scholars can use AI-enabled chatbots for information sharing, fake news verification, and other functions during an epidemic. Specifically, the AI-enabled chatbot has intelligent outbound call capability, and can actively explore and rank epidemic information, and conduct intelligent statistics, analysis, and processing.

### 5.3 Enhancing eHealth literacy

Another research avenue that can be looked into is how to improve the public’s information literacy skills facing the fact message fatigue, misinformation, and equity gaps. The concept of eHealth literacy should be included in future work, which is integrated into the concepts of health and media literacy (Norman and Skinner, 2006). Future research needs to discuss eHealth Literacy during the pandemic in three ways. First, the ability of health information acquisition from multiple information resources should be exploited (Farhan et al., 2020). Second, paying attention to good information literacy skills is essential, particularly in terms of the impacts of information literacy on appraisals of information, which can help filter overloaded information such as fake news and redundant messages (Chong et al., 2020; Lee et al., 2020). Third, developing training modules to improve eHealth literacy skills such as inviting participants to do a message deconstruction exercise. Policymakers will be suggested to encourage communities of color to engage in more social participation and dialogue through media literacy, to strengthen community capabilities, and to improve political engagement in response to information inequity (Austin et al., 2021).

From a social equity perspective, enhancing literacy on health matters should not leave behind socially disadvantaged individuals, such as people with disabilities and non-English speakers (Velasquez et al., 2020; Mein, 2020). Also, future research can propose solutions to help vulnerable populations gain equitable access to health information. Previous studies show that there is a digital divide in the contemporary information environment, and the ongoing pandemic has accentuated information disparities when many health messages are disseminated primarily on the Internet. Hence, the expansion of language services and equitable development of advanced accessible technologies would expand the availability of health networks to vulnerable populations. Moreover, it is essential to enhance the eHealth literacy of grassroots communities. People in rural areas can benefit from a community network of village health volunteers who provide information during the pandemic because they are often misinformed by social media messages. Therefore, grassroots vaccine campaigns that meet with community residents through mobile health centers or clinics at non-traditional sites like shelters will need to be examined in future research (Vicerra, 2021; Dror et al., 2021).

## 6. Conclusion

To sum up, in this research, we applied a systematic literature review to analyze individuals’ responses to COVID-19 information. The main motive of applying this methodology is the confidence that the available literature has been thoroughly and systematically searched. We found that the *Communication* discipline has the most publications on this topic. Most of the studies are primarily focused on the United States and China. Regarding the psychological responses, people’s perceptions, anxiety, fear, and conspiracy beliefs have been explored in the literature. We also combed the behavioral responses to social media and legacy media, such as wearing masks, mobility changes, and getting vaccinations. Furthermore, we found that some challenges still exist. First, during the pandemic, some health information from legacy media was not effective in containing misleading information due to the highly partisan environment. Second, COVID-19-related infodemics are exacerbating the global health crisis. Third, socially vulnerable populations are unevenly impacted by the COVID-19 information, which in turn misled their psychological and behavioral responses. Finally, we proposed a series of research agendas for future researchers.

## Data Availability

All data produced in the present work are contained in the manuscript

---

**Appendix 1.**
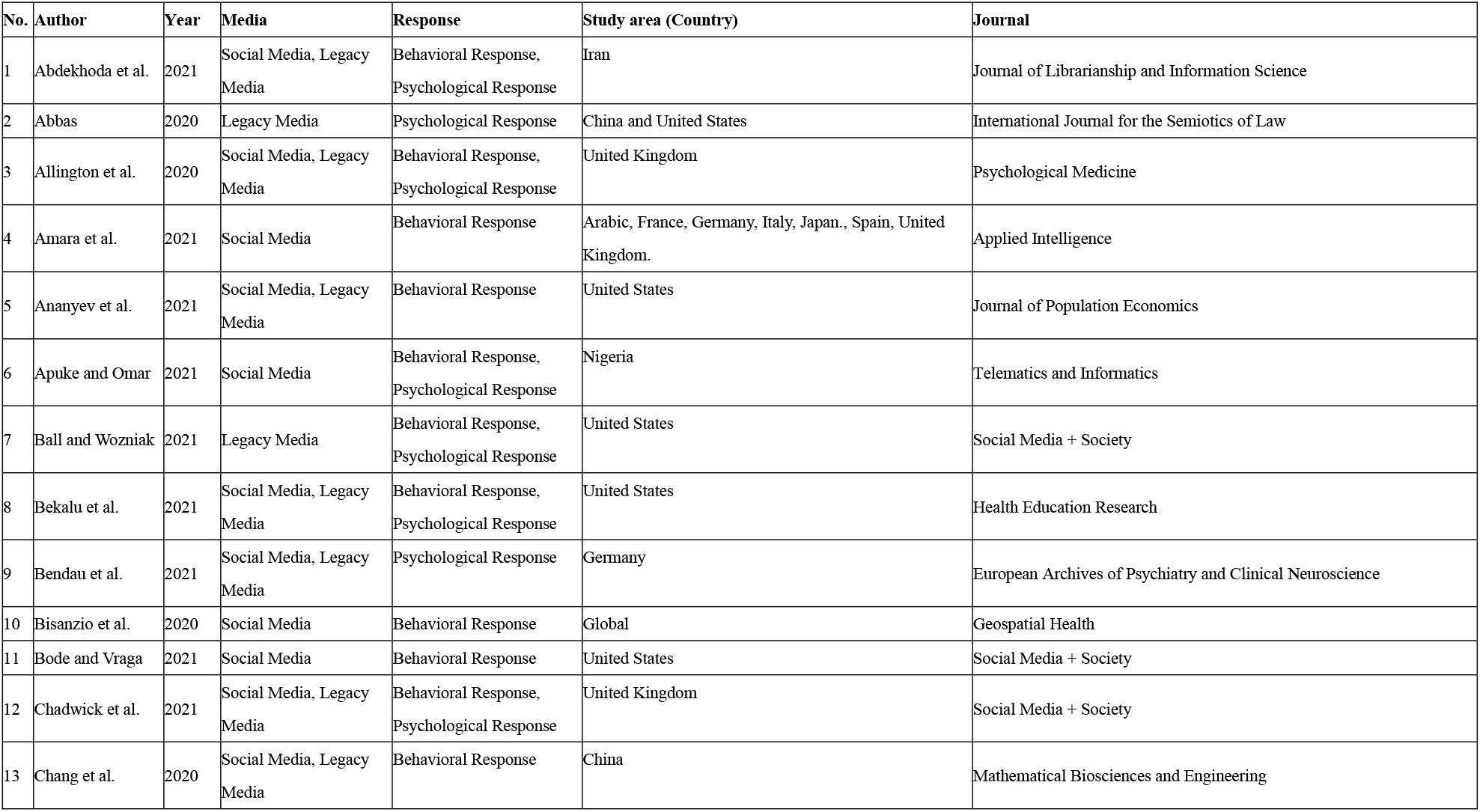
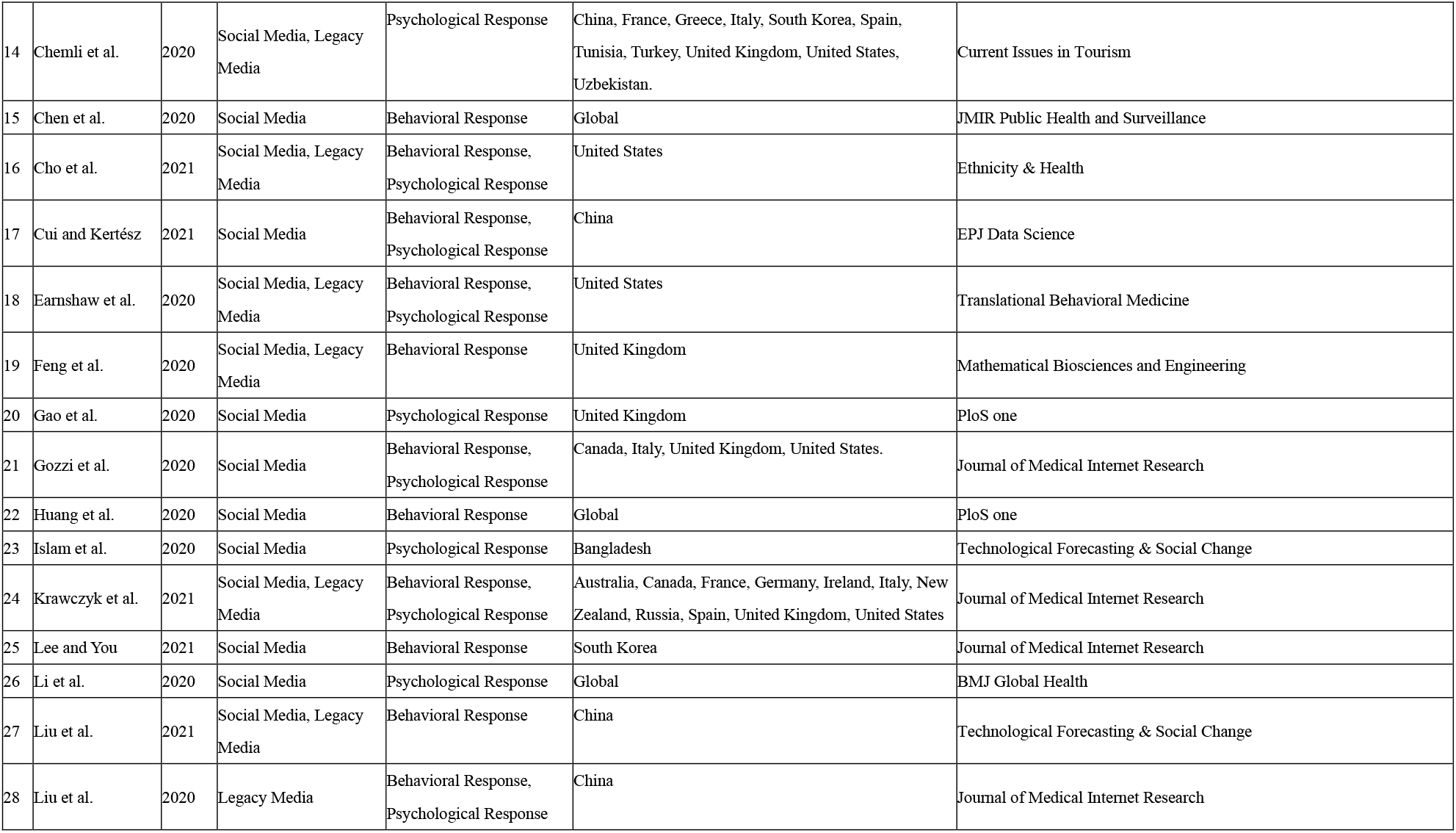
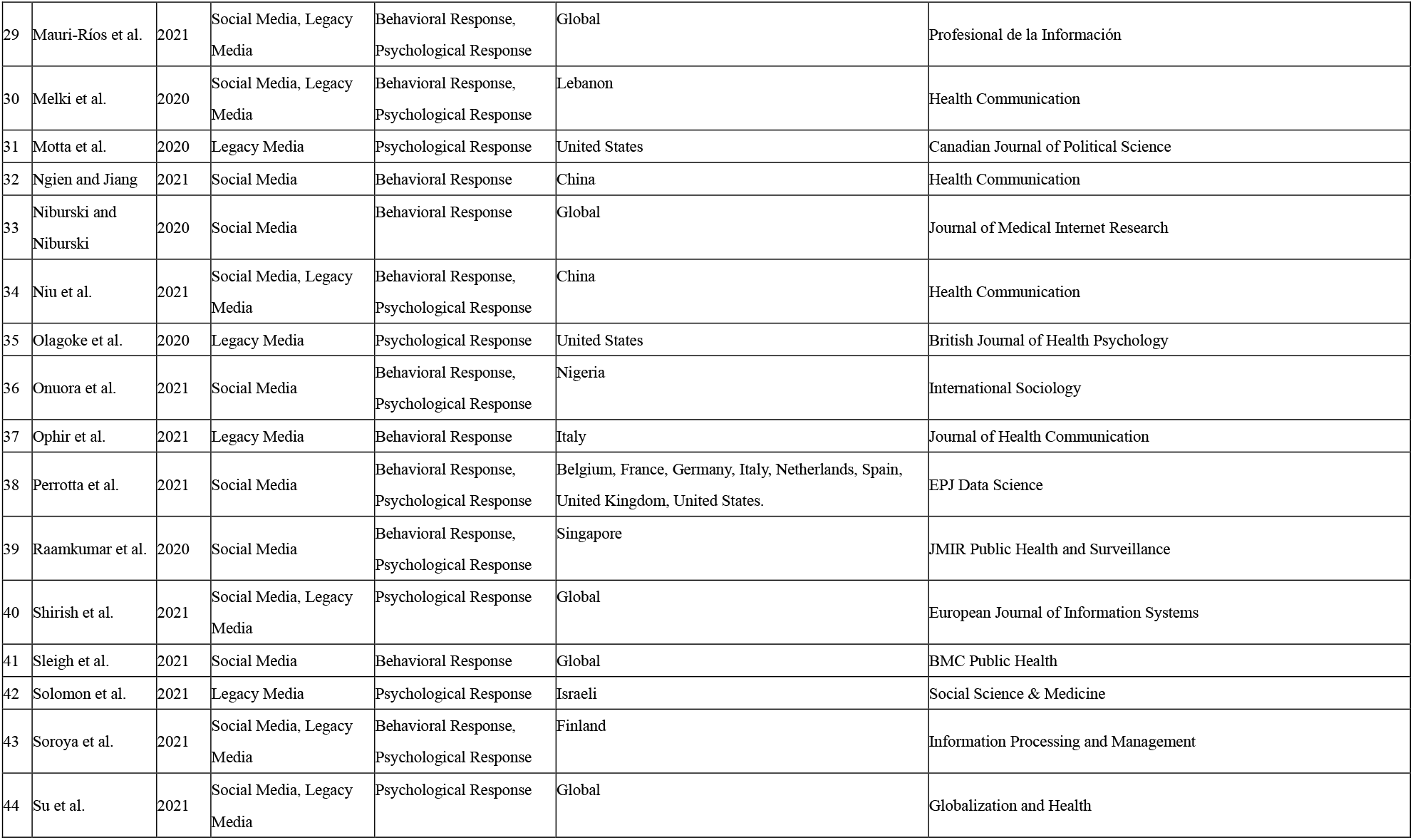
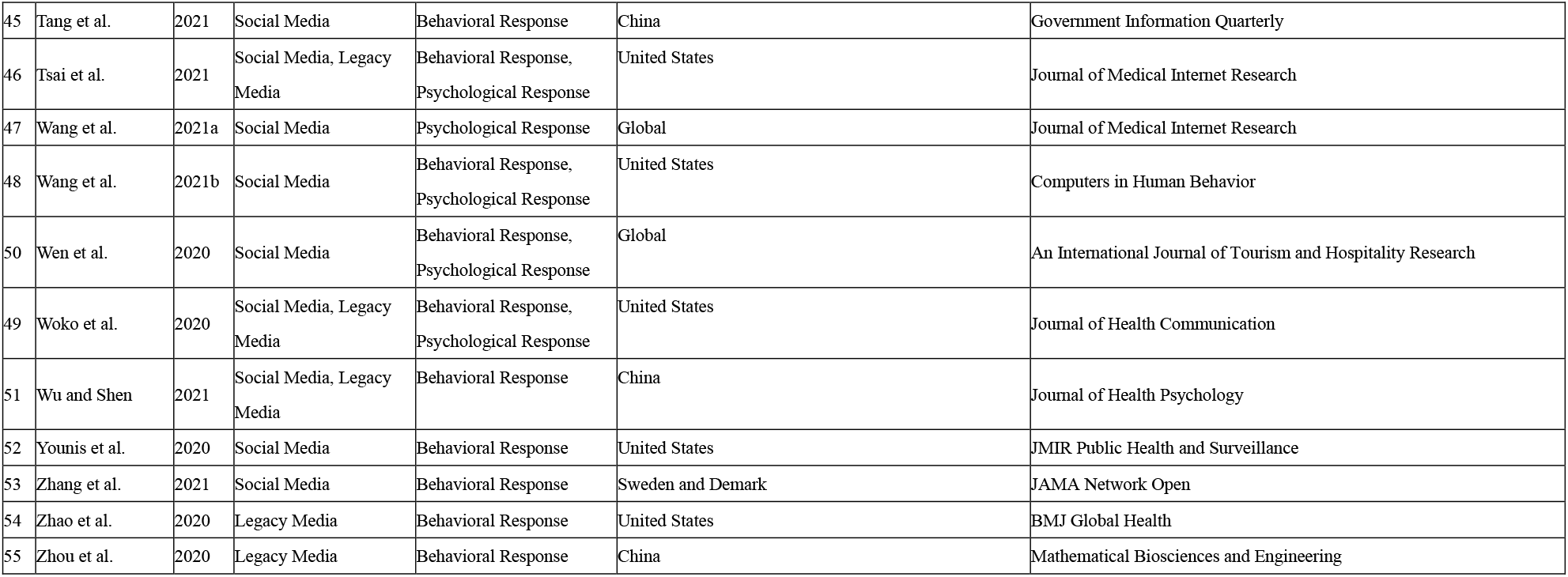

| No. | Author | Year | Media | Response | Study area (Country) | Journal |
| --- | --- | --- | --- | --- | --- | --- |
| 1 | Abdekhoda et al. | 2021 | Social Media, Legacy Media | Behavioral Response, Psychological Response | Iran | Journal of Librarianship and Information Science |
| 2 | Abbas | 2020 | Legacy Media | Psychological Response | China and United States | International Journal for the Semiotics of Law |
| 3 | Allington et al. | 2020 | Social Media, Legacy Media | Behavioral Response, Psychological Response | United Kingdom | Psychological Medicine |
| 4 | Amara et al. | 2021 | Social Media | Behavioral Response | Arabic, France, Germany, Italy, Japan., Spain, United Kingdom. | Applied Intelligence |
| 5 | Ananyev et al. | 2021 | Social Media, Legacy Media | Behavioral Response | United States | Journal of Population Economics |
| 6 | Apuke and Omar | 2021 | Social Media | Behavioral Response, Psychological Response | Nigeria | Telematics and Informatics |
| 7 | Ball and Wozniak | 2021 | Legacy Media | Behavioral Response, Psychological Response | United States | Social Media + Society |
| 8 | Bekalu et al. | 2021 | Social Media, Legacy Media | Behavioral Response, Psychological Response | United States | Health Education Research |
| 9 | Bendau et al. | 2021 | Social Media, Legacy Media | Psychological Response | Germany | European Archives of Psychiatry and Clinical Neuroscience |
| 10 | Bisanzio et al. | 2020 | Social Media | Behavioral Response | Global | Geospatial Health |
| 11 | Bode and Vraga | 2021 | Social Media | Behavioral Response | United States | Social Media + Society |
| 12 | Chadwick et al. | 2021 | Social Media, Legacy Media | Behavioral Response, Psychological Response | United Kingdom | Social Media + Society |
| 13 | Chang et al. | 2020 | Social Media, Legacy Media | Behavioral Response | China | Mathematical Biosciences and Engineering |
| 14 | Chemli et al. | 2020 | Social Media, Legacy Media | Psychological Response | China, France, Greece, Italy, South Korea, Spain, Tunisia, Turkey, United Kingdom, United States, Uzbekistan. | Current Issues in Tourism |
| 15 | Chen et al. | 2020 | Social Media | Behavioral Response | Global | JMIR Public Health and Surveillance |
| 16 | Cho et al. | 2021 | Social Media, Legacy Media | Behavioral Response, Psychological Response | United States | Ethnicity & Health |
| 17 | Cui and Kertész | 2021 | Social Media | Behavioral Response, Psychological Response | China | EPJ Data Science |
| 18 | Earnshaw et al. | 2020 | Social Media, Legacy Media | Behavioral Response, Psychological Response | United States | Translational Behavioral Medicine |
| 19 | Feng et al. | 2020 | Social Media, Legacy Media | Behavioral Response | United Kingdom | Mathematical Biosciences and Engineering |
| 20 | Gao et al. | 2020 | Social Media | Psychological Response | United Kingdom | PloS one |
| 21 | Gozzi et al. | 2020 | Social Media | Behavioral Response, Psychological Response | Canada, Italy, United Kingdom, United States. | Journal of Medical Internet Research |
| 22 | Huang et al. | 2020 | Social Media | Behavioral Response | Global | PloS one |
| 23 | Islam et al. | 2020 | Social Media | Psychological Response | Bangladesh | Technological Forecasting & Social Change |
| 24 | Krawczyk et al. | 2021 | Social Media, Legacy Media | Behavioral Response, Psychological Response | Australia, Canada, France, Germany, Ireland, Italy, New Zealand, Russia, Spain, United Kingdom, United States | Journal of Medical Internet Research |
| 25 | Lee and You | 2021 | Social Media | Behavioral Response | South Korea | Journal of Medical Internet Research |
| 26 | Li et al. | 2020 | Social Media | Psychological Response | Global | BMJ Global Health |
| 27 | Liu et al. | 2021 | Social Media, Legacy Media | Behavioral Response | China | Technological Forecasting & Social Change |
| 28 | Liu et al. | 2020 | Legacy Media | Behavioral Response, Psychological Response | China | Journal of Medical Internet Research |
| 29 | Mauri-Ríos et al. | 2021 | Social Media, Legacy Media | Behavioral Response, Psychological Response | Global | Profesional de la Información |
| 30 | Melki et al. | 2020 | Social Media, Legacy Media | Behavioral Response, Psychological Response | Lebanon | Health Communication |
| 31 | Motta et al. | 2020 | Legacy Media | Psychological Response | United States | Canadian Journal of Political Science |
| 32 | Ngien and Jiang | 2021 | Social Media | Behavioral Response | China | Health Communication |
| 33 | Niburski and Niburski | 2020 | Social Media | Behavioral Response | Global | Journal of Medical Internet Research |
| 34 | Niu et al. | 2021 | Social Media, Legacy Media | Behavioral Response, Psychological Response | China | Health Communication |
| 35 | Olagoke et al. | 2020 | Legacy Media | Psychological Response | United States | British Journal of Health Psychology |
| 36 | Onuora et al. | 2021 | Social Media | Behavioral Response, Psychological Response | Nigeria | International Sociology |
| 37 | Ophir et al. | 2021 | Legacy Media | Behavioral Response | Italy | Journal of Health Communication |
| 38 | Perrotta et al. | 2021 | Social Media | Behavioral Response, Psychological Response | Belgium, France, Germany, Italy, Netherlands, Spain, United Kingdom, United States. | EPJ Data Science |
| 39 | Raamkumar et al. | 2020 | Social Media | Behavioral Response, Psychological Response | Singapore | JMIR Public Health and Surveillance |
| 40 | Shirish et al. | 2021 | Social Media, Legacy Media | Psychological Response | Global | European Journal of Information Systems |
| 41 | Sleigh et al. | 2021 | Social Media | Behavioral Response | Global | BMC Public Health |
| 42 | Solomon et al. | 2021 | Legacy Media | Psychological Response | Israeli | Social Science & Medicine |
| 43 | Soroya et al. | 2021 | Social Media, Legacy Media | Behavioral Response, Psychological Response | Finland | Information Processing and Management |
| 44 | Su et al. | 2021 | Social Media, Legacy Media | Psychological Response | Global | Globalization and Health |
| 45 | Tang et al. | 2021 | Social Media | Behavioral Response | China | Government Information Quarterly |
| 46 | Tsai et al. | 2021 | Social Media, Legacy Media | Behavioral Response, Psychological Response | United States | Journal of Medical Internet Research |
| 47 | Wang et al. | 2021a | Social Media | Psychological Response | Global | Journal of Medical Internet Research |
| 48 | Wang et al. | 2021b | Social Media | Behavioral Response, Psychological Response | United States | Computers in Human Behavior |
| 50 | Wen et al. | 2020 | Social Media | Behavioral Response, Psychological Response | Global | An International Journal of Tourism and Hospitality Research |
| 49 | Woko et al. | 2020 | Social Media, Legacy Media | Behavioral Response, Psychological Response | United States | Journal of Health Communication |
| 51 | Wu and Shen | 2021 | Social Media, Legacy Media | Behavioral Response | China | Journal of Health Psychology |
| 52 | Younis et al. | 2020 | Social Media | Behavioral Response | United States | JMIR Public Health and Surveillance |
| 53 | Zhang et al. | 2021 | Social Media | Behavioral Response | Sweden and Demark | JAMA Network Open |
| 54 | Zhao et al. | 2020 | Legacy Media | Behavioral Response | United States | BMJ Global Health |
| 55 | Zhou et al. | 2020 | Legacy Media | Behavioral Response | China | Mathematical Biosciences and Engineering |

